# Distribution of confirmed with COVID-19 by age and gender in Mexico

**DOI:** 10.1101/2021.11.21.21264092

**Authors:** Adalberto Maldonado, Marco Reyes

## Abstract

Social, economic, and cultural factors can influence the likelihood of exposure to the virus of each person in sanitary emergencies like those of COVID-19. In this sense, parallel to the biological vulnerability to infection with SARS-CoV-2, said factors determine a complementary vulnerability to infection. Hence, they can influence in gender and age distributions of those confirmed, which is not entirely comprehended yet. The effect that age and gender can have on total vulnerability in Mexico thus far is not understood. A better understanding of such dependence can help policy optimization and decision-making to future similar emergencies. We aim to study the age and gender distributions of those confirmed with COVID-19 in Mexico. We also investigate the vulnerability to the infection with SARS-CoV-2 depending on such features. Two different samples of the Mexican population are analyzed in this non-experimental study to compare each other and evaluate the association of the result of the COVID-19 test with gender and age. Data up until the beginning of the vaccination are considered. The percentage of confirmed males is higher than females in both samples, and most tested and confirmed are working-age. Age distributions are positively skewed, with the peak in [30,39] years, which disagrees with the distribution of the Mexican population. The data suggest that the vulnerability to infection weakly depends on gender and age. Males were identified as the most vulnerable gender, and the age group [70,79] showed a higher vulnerability to infection.

## 1 Introduction

The pandemic of COVID-19, a disease produced by the new virus SARS-CoV-2 (found in December 2019, Wuhan, China), was determined as a pandemic by the World Health Organization on March 11, 2020. Since that time, the entire world has faced a complicated task to take care of the population’s health and economy (1–5).

Several features of this new virus are still unknown. The dependence by age or gender of several factors as the vulnerability to the infection of SARS-CoV-2, the likelihood that the illness becomes severe, and its mortality are not entirely understood yet; differences can be found between the results of previous works (6–11).

Sanitary emergencies like those the pandemic of COVID-19 have different effects on the population depending on gender and age. Several factors influence the experience of every person (6,8,12). The biological features of each person determine a “biological vulnerability” to the infection with SARS-CoV-2; such vulnerability measures the likelihood of infection in a person exposed to the virus. By another hand, social, economic, and cultural factors can influence the probability of exposure to the virus of each person in this kind of sanitary emergencies. In this sense, said factors determine a complementary vulnerability to the infection with SARS-CoV-2; then, these influence the gender and age distributions of those confirmed with COVID-19. For example, most strategies used to handle the pandemic include intermittent population confinement, which decreases the virus spreading (13–15). Theoretically, the likelihood of virus exposure can be strongly influenced, directly or indirectly, by gender and age since the people’s participation in confinement do it too. Some sanitary measures to face the emergency depend strongly on age; the non-working age population has a higher opportunity to participate in confinement. By another hand, the labor force participation can have significant differences by gender, which can increase the likelihood of exposure for some (primarily in risk activities).

The number of databases that include categories, such as age, gender, comorbidities, education level, ethnicity, etcetera, is limited and less helpful if collected separately (6,7,16). More complete databases allow data breakdowns to find trends or patterns that can help policy optimization and decision-making. However, not be overlooked that multiple factors make it difficult to obtain and analyze data, which can lead to skewed or confusing results (6,7). The understanding of the pandemic can be improved by taking advantage of the large databases available from different countries. It is not yet clear the effect that gender and age have on vulnerability to infection (6–8,10,12,16–18), for example. In Mexico, the gender distribution of those confirmed as of December 24, 2020, seems uniform (50.5% males and 49.5% females). However, several features which can influence gender distribution are missing out in such perception. The Mexican population is composed mainly of females (51.2% (19)), but the labor force participation rate is higher in males (19), so the likelihood of virus exposure could be significantly different between genders. In this work, we analyzed the age and gender distributions of those confirmed with COVID-19 in two different samples of the Mexican population; both databases include age and gender. We also evaluate the effect that such features have on vulnerability to the infection with SARS-CoV-2. We use data up until the day of the beginning of vaccination in Mexico (20).

## 2 Method

In the research, are considered two databases. Sample 1 is the open official data from the Ministry of Health (21), which comes from the sentinel surveillance model applied in Mexico. Said model consists of the collection, integration, verification, and analysis of detailed epidemiological information from a small set of health monitoring units (22), which is a limitation of the model.

Sample 2 was provided by BMSA Group, a Mexican business group that provides health solutions for business entities that unwrap in Mexico. Their data comes from the tests applied as a part of their strategy to handle the emergency in the companies attended; hence, most of the tested people are working age. The strategy reported by the business group includes contact tracing, periodic control, filter on admission, and application of sanitary measures, which may lead to multi-tested people. Duplicates in the database were eliminated to avoid multiple counting. The samples were collected by qualified workers of *BMSA Group* and analyzed by *Orthin* laboratory or *Laboratorio Imagen*. The companies attended by *BMSA Group* (which include entities with an international presence) belong to different sectors and make activities considered essential, so they did not suspend activities during the pandemic. Therefore, their population could not participate in the confinement, which increased its exposure risk to SARS-CoV-2.

The proportions by age, and gender, of confirmed, negative, and tested were analyzed and compared with the total Mexican population to determine if the pandemic depends on such features. We use data until December 24, 2020, the day of the beginning of vaccination in Mexico (20), to avoid its effect on results.

## 3 Results

As of December 24, 2020, in Mexico 3,277,138 tests were applied, and 1,445,624 infected were detected. The proportion of males in the analyzed population is 48.2% and 50.5% in confirmed cases.

The tested and confirmed age distributions are not uniform in sample 1. Both are distributions with positive skewness (Figures 1-(a)-(b)). The peak of the distributions is in the age range from 30 to 39 years. There concentrate 22.8% of the tests applied and 21.8% of those confirmed. As seen in Table 1, the age distributions from males and females are analogous to those of the total population.

**Figure 1.**
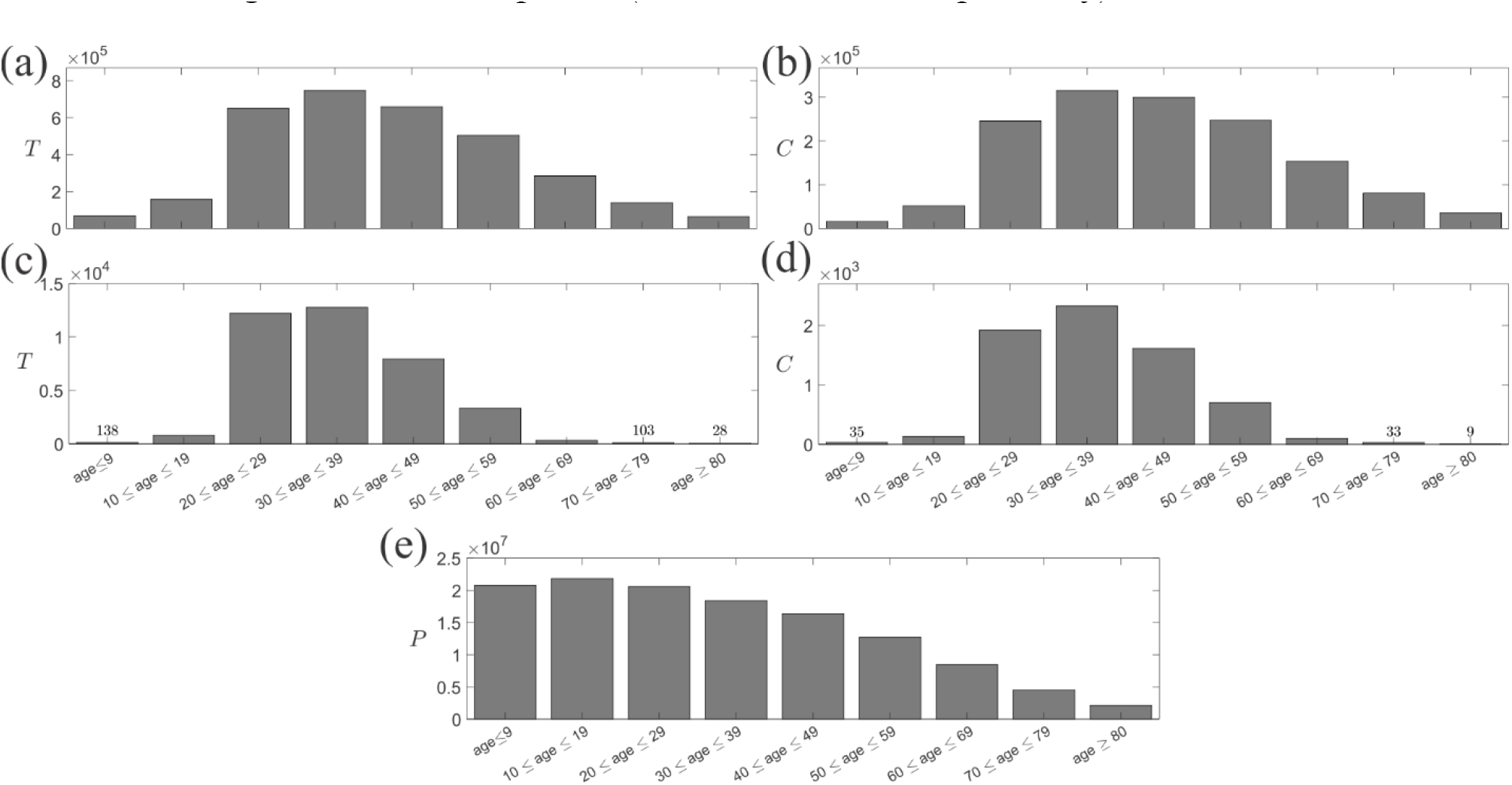
Age distributions as of December 24, 2020, for (a) Applied tests (T) in Mexico, (b) Confirmed cases (C) in Mexico, (c) Applied tests (T) by BMSA Group, (d) Confirmed cases (C) by BMSA, and (e) Mexican population.

**Table 1.** The number of (a) tested and (b) confirmed in every age group for males, females, and the total population in the official database.

| (a) | ≤ 9 | 10-19 | 20-29 | 30-39 | 40-49 | 50-59 | 60-69 | 70-79 | ≥ 80 |
| --- | --- | --- | --- | --- | --- | --- | --- | --- | --- |
| Total | 68996 | 157511 | 650378 | 747304 | 659651 | 505088 | 284958 | 139730 | 63522 |
| Males | 36653 | 76224 | 305520 | 354981 | 310678 | 241550 | 146004 | 74200 | 32583 |
| Females | 32343 | 81287 | 344858 | 392323 | 348973 | 263538 | 138954 | 65530 | 30939 |
| (b) | ≤ 9 | 10-19 | 20-29 | 30-39 | 40-49 | 50-59 | 60-69 | 70-79 | ≥ 80 |
| Total | 16102 | 51848 | 245705 | 315298 | 299625 | 247025 | 153771 | 80659 | 35591 |
| Males | 8492 | 25238 | 117935 | 156760 | 149908 | 125767 | 82287 | 44695 | 19309 |
| Females | 7610 | 26610 | 127770 | 158538 | 149717 | 121258 | 71484 | 35964 | 16282 |

By another way, through 44,420 tests, as of December 24, 2020, BMSA Group had detected 7,637 cases of COVID-19 in the attended population. In this sample 2, most of the tests and confirmed cases are from males (65.0% and 67.4%, respectively); there is also a small group of cases whose gender was not reported (10.9% and 5.6%, respectively).

As with the official data (sample 1), the distribution of confirmed cases by age in sample 2 is neither uniform nor Gaussian; it shows positive skewness (Figure 1-(d)). The age range with the highest number of cases is 30 to 39 years (33.8%), and where less, in the tails of age distribution, as can be seen in Table 2-(b), where the confirmed by age are shown. The confirmed between 20 and 29 years old, and 40 and 49 years old, also represent a significant percentage. The age distribution of confirmed cases of each gender is similar to those of the total confirmed cases (and each other), as seen in Table 2-(b).

**Table 2.** The number of (a) tested and (b) confirmed in every age group for males, females, and the total population in the database of BMSA Group. The numbers from the population whose age was not reported (NR) are also shown.

| (a) | $\leq 9$ | 10-19 | 20-29 | 30-39 | 40-49 | 50-59 | 60-69 | 70-79 | $\geq 80$ | NR |
| --- | --- | --- | --- | --- | --- | --- | --- | --- | --- | --- |
| Total | 138 | 783 | 12195 | 12770 | 7946 | 3329 | 313 | 103 | 28 | 6815 |
| Males | 73 | 482 | 8428 | 8911 | 5668 | 2529 | 162 | 48 | 13 | 2541 |
| Females | 64 | 287 | 3361 | 3489 | 2113 | 771 | 151 | 55 | 15 | 423 |
| (b) | $\leq 9$ | 10-19 | 20-29 | 30-39 | 40-49 | 50-59 | 60-69 | 70-79 | $\geq 80$ | NR |
| Total | 35 | 133 | 1923 | 2326 | 1615 | 703 | 98 | 33 | 9 | 762 |
| Males | 23 | 74 | 1371 | 1635 | 1118 | 524 | 51 | 16 | 4 | 335 |
| Females | 12 | 59 | 533 | 667 | 485 | 178 | 47 | 17 | 5 | 56 |

The number of confirmed (total and by gender), with lacking age within the database, is higher than those found in tails of the distribution (Table 2-(b)). Furthermore, the number of confirmed cases with missing gender and age (371) in the database is significantly higher than the confirmed cases with only the gender not reported in the different age groups (≤24).

Also, the applied tests distribute by age groups in different percentages (Figure 1-(c)). Regardless of the gender, the age distribution of the tested is analogous to those of confirmed cases (Table 2); however, the skewness is more apparent for tests, as shown in Figures 1-(c)-(d).

## 4 Discussion

The tested population, which includes infected (confirmed) and not infected (negative) people with and without symptoms, is a sample of the total population biased towards infected cases since they were selected for suspected infection; in this sense, said population has a large proportion of those exposed to the virus. This selection procedure of the tested population does not influence a priori the gender or age distributions; however, bias in selection can exist (confinement limits the exposure for tails of age distribution, for example), which can influence the accuracy of interpretations in this non-experimental study. The study has the natural limitations of non-experimental studies; there was no control over the variables involved (like the virus exposure, comorbidities, health status, etcetera, of the addressed population). In this sense, the study does not have high accuracy.

The fact that the highest number of confirmed cases in sample 2 are from males does not necessarily imply that they are more vulnerable to SARS-CoV-2; the population of the set of companies is predominantly males. Data from the Mexican population also show that number of confirmed males is higher than those of females, no matter the total population is composed mainly of females (19). Using the chi-square test, we evaluate the differences in the proportions by age and gender that the two samples (or different subsets) have with the Mexican population to determine their representativity. In the cases representative of the Mexican population, we evaluate the relation of test result (confirmed or negative) with gender (and age) by the chi-square test of independence. We interpret the dependence between those variables as dependence in vulnerability to SARS-CoV-2 infection, a vulnerability that includes several factors such as the social, biological, economic, and cultural, by example. Hence, independence between such variables means independence in the vulnerability to SARS-CoV-2 infection.

### 4.1 Analysis by gender

For sample 1, the difference in proportions by gender of applied tests (48.2% of males and 51.8% of females (21)) with the total population (48.8% of males and 51.2% of females (19)) is not statistically significant 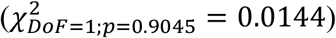^1^, which shows that, relative to gender, the tested population is a representative sample of the Mexican population. The same result appears in every age group until 79 years old 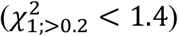; for the tested with 80 years old and higher 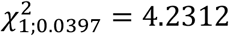, hence such age group is not representative of the Mexican population.

The chi-square test of independence applied to the total tested population (and every age group in [20,79]) shows that result of the COVID-19 test is related to gender; the power of the test is high (≈ 1)^2^ and 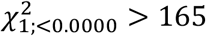, which suggests that vulnerability to SARS-CoV-2 infection depends on gender. The found dependence of gender is consistent with discussed and the results in several previous related studies (6–8,10,12,16,18) and with outbreaks of a similar virus (23). The size effect evaluated with Cramer’s *V* is below 0.1, so the dependence is weak. We interpret the positive test rate (*T*_+_) as a measure of vulnerability. The positive test rate is higher for males in all these cases (Figure 2); therefore, the vulnerability of males is higher than that of females, which agrees with the reported by Gudbjartsson et al. (10) for the Icelandic population.

**Figure 2.**
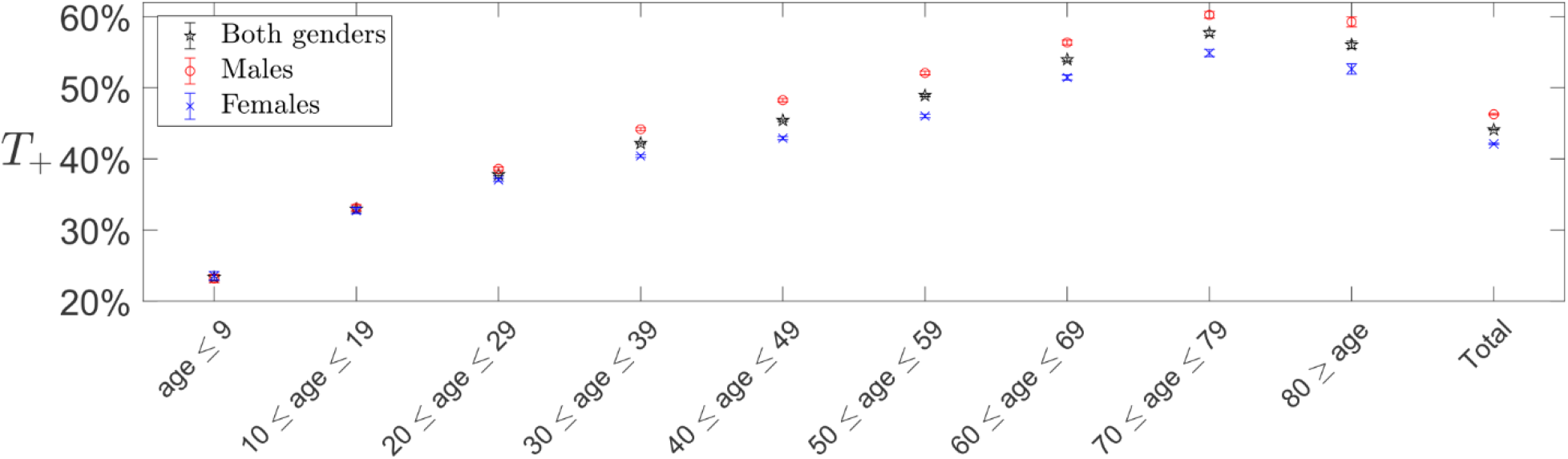
Positive tests rate (T_+_) for every age group and the total, and by gender. The 99% confidence interval is also shown.

In contrast, for the age groups with those tested up until 19 years old 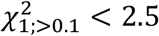; however, the probability of a type II error is high (*β* > 0.6) in such groups. In addition, the confidence intervals of *T*_+_ overlap each other for age groups in [0,19] years (Figure 2), which leads to the not significant differences by gender. Therefore, we attribute the discrepancy to the number of the data in such age intervals, which appears to be insufficient for a non-experimental study.

For sample 2, the differences in gender proportions with the total Mexican population are statistically significant (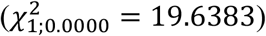), as in age groups within 10 and 59 years old and those of age not reported (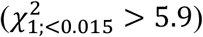). In the rest of the age groups, the difference between proportions is not significant (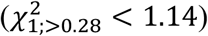), so, relative to gender, these are representative samples of the respective age group of the general Mexican population. However, the sample size in such age groups is small, leading to a very high probability of a type II error (*β*); *β* > 0.94 for age groups where people have 60 years and older, while *β* > 0.59 for kids up until 9 years old. Hence, sample 2 does not allow have conclusions about the gender dependence in vulnerability to infection with SARS-CoV-2.

### 4.2 Analysis by age

As suggested in Figures 1-(a) and (e), significant differences exist in the age proportions of the total population if compared to those tested of sample 1 (for males, females, and no matter the gender 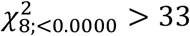), showing that relative to age, tested population is not a representative sample of the Mexican population. However, by removing the age groups in [0,19] years, the differences in such proportions become statistically not significant (for males, females, and no matter the gender 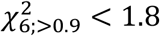, which shows that, relative to age, tested population with 20 years and older is a representative sample of the Mexican population. The chi-square test of independence applied to tested with 20 years and bigger has high power (≈ 1) and showed that no matter the gender, the result of the COVID-19 test is weakly related with age (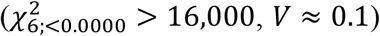), suggesting that vulnerability to SARS-CoV-2 infection also depends on age. The existence of association is consistent with the reported in earlier related works (10,17,18). In the age group [0,9], *T*_+_ is lower than those rest of the age groups, which agrees with reported for the Icelandic population (10). The positive test rate grows with age up until age group [70,79] no matter the gender, the group where *T*_+_ reaches its highest value; hence such age group is the most vulnerable.

The population in age group [0,19] years, where there exist a high proportion of students, has higher possibilities to participate in the confinement, decreasing its likelihood of virus exposure. The lower probability of exposure joined to the strategy applied in Mexico explains the high difference between those tested and the Mexican population for age groups [0,9] and [10,19]. As a result, we cannot make conclusions about the age dependence in vulnerability to infection with SARS-CoV-2 for minors.

Relative to age, sample 2 is not a representative sample of the Mexican population. The differences between the age proportions of the total population and those tested are statistically significant regardless of gender (for the males, females, and no matter the 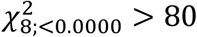), even removing the age groups with the minors (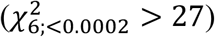). Therefore, no conclusions can do about the age dependence in vulnerability to SARS-CoV-2 infection. Most of those attended population by BMSA Group is on working-age, which influences the age distribution. The tests applied to the non-working age population were carried out only when they live with a confirmed case that belongs to the labor force of the companies attended; hence the number of tests applied in the distribution’s tails is much lower.

## 5 Conclusions

Two different samples of the Mexican population were analyzed in this non-experimental work to study the age and gender distributions and investigate if vulnerability to infection depends on such features. The gender proportions of confirmed cases in sample 1 are similar (50.5% of males and 49.5% of females), while in the other sample, 67.4% of confirmed with COVID-19 are males, 38.2% are females, and in the remaining 5.6%, the gender was missing. The age distribution of those tested and confirmed have positive skewness for both samples, which disagrees with the distribution of the Mexican population; the age group with the highest number of confirmed and tested is [30,39].

Some data allowed us investigating the vulnerability to SARS-CoV-2 infection, which is influenced by several factors, as social, biological, economic, and cultural. Analysis suggests that vulnerability to SARS-CoV-2 infection has a dependence on gender and age, which is consistent with previous related works (6–8,10,12,16–18); however, the found association is weak. We identified the males as the most vulnerable gender and the population aged between 70 and 79 years old as the age group with the higher vulnerability (no matter the gender).

## Data Availability

The data by age and gender supporting the findings of this study are available from http://dx.doi.org/10.17632/6jgry5n75w.1.

https://www.gob.mx/salud/documentos/datos-abiertos-152127

## Declaration of Competing Interest

The authors declare the following financial interests/personal relationships that may be considered as potential competing interests. A. Maldonado is the director of the data science division in BMSA Group, the company that financed the study and provided sample 2. M. Reyes participated in the development of this work under his work activities within the data science division of BMSA Group.

## Acknowledgments

The authors express their gratitude to Dr. J. Cabañas for his comments and support.

## Data sharing

The data by age and gender supporting the findings of this study are available from http://dx.doi.org/10.17632/6jgry5n75w.1. Actualized data of sample 1 are available in Ref. (21).

## Footnotes

1 Where *DOF* is the number of degrees of freedom and *p* is the p-value.

2 For those under 9 years old, the power is around 0.8.

## Notes

### Funding Statement

This work was supported by BMSA Group, a Mexican company that did not influence the results at all.

### Author Declarations

The Faculty of Medicine's Research Ethics Committee of the National Autonomous University of Mexico stated that the study is classified as risk-free research and does not require ethical oversight. The letter with the resolution of the ethics committee in English is in process and, upon your request, can be sent when it is available.

